# MB-CasFire™: A Plug-and-Play, Thermostable CRISPR Enzyme for Precise Nucleic Acid Detection

**DOI:** 10.64898/2026.11.06.26367472

**Authors:** James P. Broughton, Emel Alpay, Nick J. Fantin, Sara Ansaloni, Sarah Elsener, Jaeyoung Kirsten Jung, Yining Zhang, Isaac Z. Madrid, Stacey A. Shiigi, Srijay Rajan, Ishita Jain, William Douglass Wright, Carley G. Hendriks, Stepan Tymoshenko, Vlada Ratushna, Sierra Lee, Mavis Warner, Ryan Hong, Xinyi Sui, Shruti Panchal, Adam L. Garske, Xin Miao, Janice S. Chen

**Affiliations:** Mammoth Biosciences, Inc., 1000 Marina Blvd, Suite 600, Brisbane, CA 94005, USA

## Abstract

CRISPR-based detection technologies have the potential to enable precise, rapid, and cost-effective diagnostic and biosurveillance solutions by pairing sensitive and specific nucleic acid detection with a simple workflow. Combining isothermal amplification and CRISPR detection in a single reaction as a “one-pot” detection method can meet these needs, but only if the CRISPR enzyme remains active at the temperatures and salt concentrations that isothermal amplification requires. Here we describe MB-CasFire™, a CRISPR-Cas enzyme engineered from a naturally thermostable Cas12a for improved stability and performance, as a “plug-and-play” DETECTR^®^ (DNA Endonuclease-Targeted CRISPR Trans Reporter) enzyme for sensitive and specific nucleic acid detection. MB-CasFire remains active at elevated temperatures in amplification buffers and supports one-pot detection with loop-mediated isothermal amplification (LAMP) and nicking enzyme amplification reaction (NEAR), eliminating transfers between amplification and readout, and increasing the signal-to-noise to enable precise nucleic acid detection. This method, called OnePot DETECTR, is also compatible with commercially available isothermal master mixes, including those from New England Biolabs, allowing assays to be assembled from standard reagents. Using MB-CasFire, we detected SARS-CoV-2 in nasal swab samples with a LAMP-based OnePot DETECTR assay and *Neisseria gonorrhoeae* in urine samples with a NEAR-based OnePot DETECTR assay. MB-CasFire also functioned in multiplex detection and single-nucleotide polymorphism discrimination assays for antibiotic resistance genotyping. These results support MB-CasFire as a versatile detection enzyme that simplifies assay architecture for CRISPR-based detection assays that amplify, recognize, and report the presence of nucleic acid targets in a single reaction without thermocycling.

## INTRODUCTION

The unexpected finding that certain CRISPR-Cas12a proteins unleash indiscriminate single-stranded DNase activity led to the development of DETECTR^®^ (DNA Endonuclease-Targeted CRISPR Trans Reporter), a method that couples programmable nucleic acid recognition to enzymatic signal generation. In Cas12a-mediated detection assays, guide RNA (gRNA)-directed target recognition activates trans-cleavage (also known as collateral cleavage) of single-stranded DNA (ssDNA), allowing activation of a fluorescence signal from a ssDNA reporter (Chen et al., 2018). However, trans-nuclease activity by Cas12a alone often provides insufficient sensitivity to detect low-abundance diagnostic targets (Huyke et al., 2022). Although established isothermal amplification methods such as loop-mediated isothermal amplification (LAMP), recombinase polymerase amplification (RPA), and nicking enzyme amplification reaction (NEAR) can achieve high sensitivity (Notomi et al., 2000; Piepenburg et al., 2006; Nie et al., 2014), false positive signals remain a concern due to non-specific amplification that can be detected when using intercalating dyes such as SYBR Green I (Piepenburg et al., 2006; Bhadra et al., 2021). Unlike sequence-specific probes such as molecular beacons which rely only on sequence complementarity, CRISPR-based detection requires both gRNA-directed target recognition and enzyme activation before reporter cleavage can occur, adding an extra layer of specificity to reduce false positives while also enabling detection of single-nucleotide polymorphisms (SNPs) and new assay formats. This capability motivated the development of the first-generation DETECTR method, which combined Cas12a trans-cleavage activity with isothermal amplification to increase target abundance and specificity as a “two-pot” workflow without thermal cycling (Chen et al., 2018).

Running amplification and CRISPR detection simultaneously in a single “one-pot” reaction mixture further simplifies the workflow by removing post-amplification transfers and reducing opportunities for amplicon carryover (Ding et al., 2020). The integration of CRISPR-based detection with isothermal amplification in a single reaction requires compatible reaction temperatures, buffer conditions, and enzyme activities. Many isothermal amplification methods function at temperatures above 55°C, creating a need for a Cas enzyme that can bind target DNA and perform trans-nuclease activity at these temperatures. Thermostable Cas12a orthologs and engineered Cas12a and Cas12b variants have enabled concurrent LAMP-based detection, establishing both natural diversity and protein engineering as routes to thermal compatibility (Pena et al., 2023; Nguyen et al., 2023; Pan et al., 2025). Despite these examples in the literature, many native Cas enzymes remain challenging to use in isothermal amplification reactions because they must also avoid interfering with target amplification, and resist inhibition by amplification reagents, such as high salt and dNTP concentrations (Tong et al., 2024; Pan et al., 2025). Similarly, commercial availability and prohibitively long gRNAs can limit access to some thermostable Cas enzymes. The limitations of current Cas enzymes motivates discovery and engineering of nucleases specifically for diagnostic applications.

Sequence discrimination introduces a further requirement when assays must distinguish alleles differing by a single nucleotide. Cas12a is sensitive to single-nucleotide mismatches between the gRNA and target, particularly when the mismatch occurs in the seed region adjacent to the protospacer-adjacent motif (PAM) (Swarts et al., 2017). However, successful design of allele-specific gRNAs depends on the position of the mismatch and sequence context (Mao et al., 2024). Cas12a-based genotyping has demonstrated SNP detection, including identification of SARS-CoV-2 variants (Fasching et al., 2022). SNP detection has also been achieved by coupling Cas12b detection with NEAR (Bai et al., 2025). Single-nucleotide discrimination therefore adds an important capability to integrated diagnostic assays, complementing thermal compatibility and reporter-cleavage activity.

Here, we describe MB-CasFire™, a thermostable Cas12a engineered for diagnostic applications. MB-CasFire supports simultaneous isothermal target amplification and fluorescence detection through trans-nuclease activity, a method called OnePot DETECTR which can be applied to multiple isothermal amplification approaches including LAMP and NEAR. We also demonstrate MB-CasFire’s capacity for single-nucleotide discrimination, extending its utility from target detection to analysis of sequence variation. Alongside MB-CasFire, we describe two additional engineered Type V effectors, MB-CasDx8 and MB-CasDx9 that are compatible with isothermal amplification. With MB-CasFire as the principal focus, this study examines the applications of engineered Cas enzymes as a basis for the next generation of CRISPR-based diagnostics.

## RESULTS

To establish a OnePot DETECTR assay, we combined isothermal target amplification and CRISPR-mediated trans-cleavage of a fluorescent reporter DNA molecule in a single reaction maintained at a constant temperature (Fig. 1a). We first evaluated EnGen LbaCas12a (NEB) in an RT-LAMP reaction to detect SARS-CoV-2 RNA, monitoring amplification with a fluorescent DNA-binding dye and trans-nuclease activity with a fluorescent ssDNA reporter. The amplification-associated fluorescence from the DNA-binding dye increased strongly, whereas the LbaCas12a reporter signal was slightly elevated, but remained near baseline throughout the 60 min reaction (Fig. 1b). The disparity between amplification as detected by the DNA-binding dye and the ssDNA reporter motivated us to identify a nuclease with greater activity in one-pot reaction conditions.

**Figure 1.**
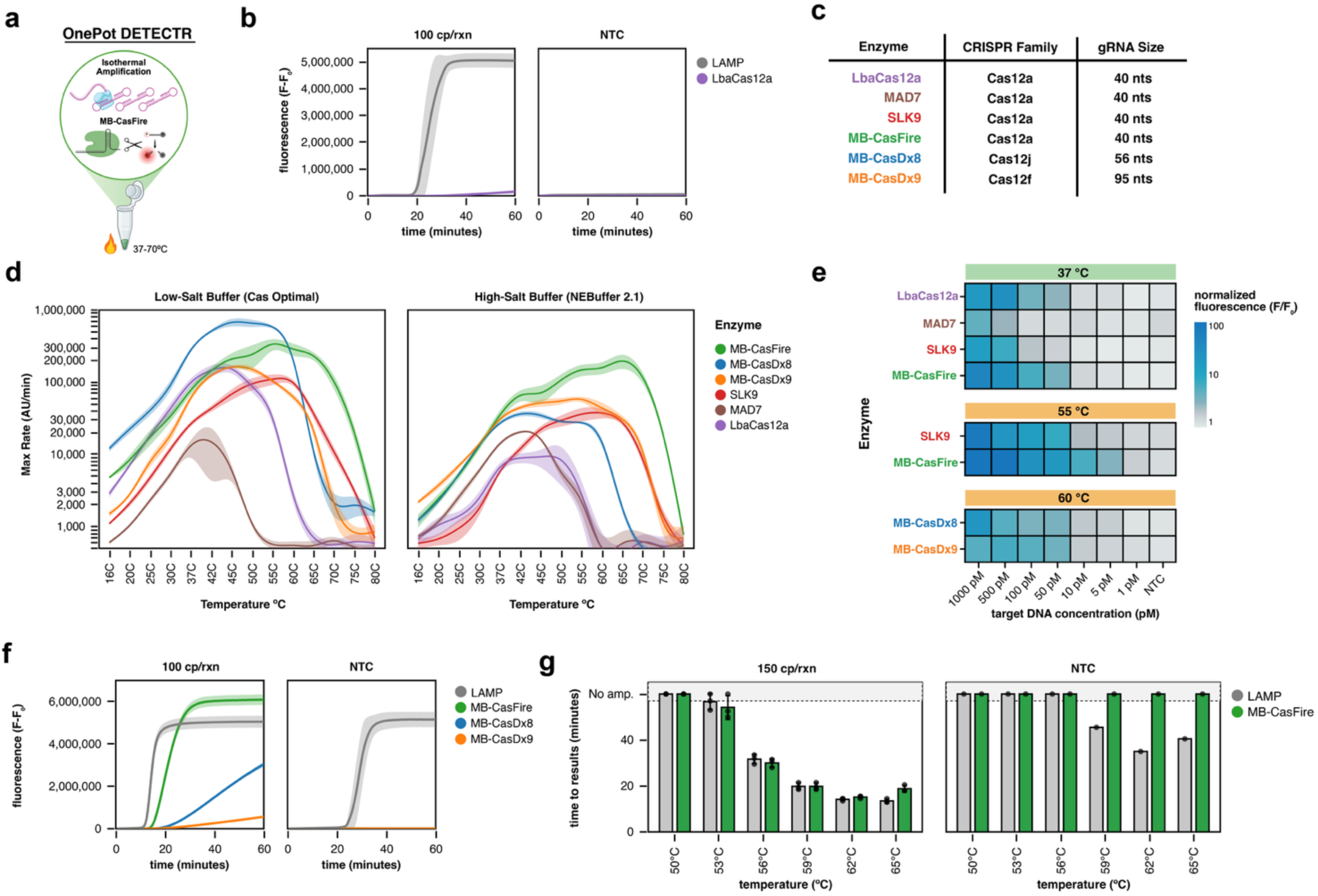
Characterization of engineered CasDx enzymes for OnePot DETECTR. **(a)** Schematic showing the generalized format of OnePot DETECTR reactions. The assay temperature is dependent on the type of isothermal reaction that is coupled to CRISPR-based detection. Created in BioRender. Broughton, J. (2026) https://BioRender.com/wp5o48q. **(b)** OnePot DETECTR using RT-LAMP to detect SARS-CoV-2 RNA with EnGen LbaCas12a (NEB) at 56°C shows weak signal (purple) from the Cas enzyme compared to the signal from the DNA-binding dye (gray). Shaded areas represent ±1 SD. n=4. **(c)** List of enzymes, their predicted Cas family based on sequence homology, and guide RNA length in nucleotides (nt). **(d)** Temperature dependence of trans-nuclease activity for six Cas12 enzymes in low-salt buffer (Cas optimal) and high-salt buffer (NEBuffer 2.1). Maximum reaction rate (AU/min), taken as the steepest slope of the reporter fluorescence trace, is plotted against incubation temperature. Shaded areas represent ±1 SD. n=4. **(e)** Analytical sensitivity of Cas enzymes at 37, 55 and 60°C in low-salt buffer (Cas optimal). Heat map of normalized reporter fluorescence (F/F_0_) after 60 minutes at the indicated temperature across a range of target DNA concentrations. n=3. **(f)** RT-LAMP-based OnePot DETECTR performance at 53°C across three engineered CasDx enzymes for the detection of influenza B virus RNA. Shaded areas represent ±1 SD. n=3. **(g)** RT-LAMP-based OnePot DETECTR performance for SARS-CoV-2 RNA at 150 cp/rxn and in NTCs across 50-65°C. LAMP amplification (gray bars) is monitored by a DNA-binding dye, and MB-CasFire performance (green bars) is monitored by ssDNA cleavage reporter fluorescence. Time to result (TTR) is plotted for each readout; the upper category denotes reactions without a detected result during the 60 minute assay window. Error bars represent ±1 SD. n=3.

To identify CRISPR-Cas enzymes with potentially advantageous properties for OnePot DETECTR, we built profile hidden Markov models (HMMs) for dozens of Cas-like protein families. We used these profiles to search our metagenomic database of more than 35 billion protein sequences, some sourced from extreme environments, with HMMER (Eddy, 2011). This search identified hundreds of previously uncharacterized Cas proteins across Type V families, which we expressed in *E. coli* and screened as lysates for trans-nuclease activity. Among the enzymes that showed trans-nuclease activity, the most active was a naturally thermostable Cas12a, which we selected as the starting point for MB-CasFire. We then engineered this enzyme for greater thermostability and for compatibility with isothermal amplification buffers. Using a DETECTR-based PAM profiling assay, we determined that MB-CasFire was nearly PAM-less (Supplementary Fig. 1). We also identified mesophilic enzymes with modest trans-nuclease activity, including MB-CasDx8 (a Cas12j enzyme) and MB-CasDx9 (a Cas12f enzyme), and asked whether they could also be engineered to improve thermostability and salt tolerance. Four rounds of protein engineering increased the activity and extended the operating temperature ranges for each enzyme in an isothermal amplification buffer. For MB-CasDx8, protein engineering converted an enzyme with little activity above 50°C into one with substantial activity at 55-60°C (Supplementary Fig. 2a). For MB-CasDx9, the final variant exhibited strong activity across 45-60°C and retained activity at 65°C (Supplementary Fig. 2b). These engineering efforts provided three candidates for subsequent comparison under conditions relevant to OnePot DETECTR.

To compare MB-CasFire with other Type V enzymes, we tested it alongside EnGen LbaCas12a (NEB), MAD7 (Horizon Discovery; Liu et al., 2020), SLK9 (Sherlock Biosciences; Pena et al., 2023), MB-CasDx8, and MB-CasDx9 (Fig. 1c). We measured trans-nuclease rates from 16 to 80°C in two buffers, a low-salt reaction buffer that favors Cas enzyme activity and a high-salt buffer (NEBuffer 2.1) chosen to approximate the ionic conditions of isothermal amplification. In the low-salt buffer, MB-CasDx8 had the highest peak activity of the six enzymes, at approximately 45-50°C, whereas MB-CasFire peaked at 55°C. Activity was lower in the high-salt buffer across all tested enzymes, but MB-CasFire had the highest trans-nuclease rate from 50 to 75°C (Fig. 1d), suggesting it may be a promising candidate for OnePot DETECTR assay development.

We next examined the sensitivity of the enzymes at their optimal temperature in a low-salt buffer across a range of dsDNA target concentrations. MB-CasFire demonstrated the best sensitivity of the enzymes at 55°C down to a target concentration of 5 pM. SLK9, MB-CasDx8, and MB-CasDx9 exhibited similar sensitivity at 50 pM (Fig. 1e). To determine how these activities translated to an amplification-coupled assay, we compared the three engineered enzymes in OnePot DETECTR to detect influenza B virus RNA across a range of temperatures. Although MB-CasDx8 and MB-CasDx9 were compatible with the one-pot format below 59°C, MB-CasFire produced the strongest reporter signal among the three enzymes and the least apparent delay relative to the LAMP dye signal (Fig. 1f, Supplementary Fig. 2c). This result supported the selection of MB-CasFire for subsequent assay development.

To define MB-CasFire’s functional temperature range, we tested a OnePot DETECTR assay for SARS-CoV-2 RNA from 50 to 65°C. Reactions used NEB’s WarmStart^®^ Multi-Purpose LAMP/RT-LAMP 2X Master Mix (NEB, M1708) and contained both a DNA-binding dye to monitor amplification and a fluorescent ssDNA reporter to monitor MB-CasFire trans-nuclease activity (Fig. 1g). Detection times were similar for the two signals, with MB-CasFire generally lagging amplification by a couple of minutes, consistent with the need for amplicon to accumulate before Cas detection (Supplementary Fig. 2d). In some no-template control (NTC) reactions, however, the DNA-binding dye produced a signal indicative of non-specific amplification, whereas the MB-CasFire reporter remained negative at every temperature tested. These results underscore the specificity advantage of having a second recognition layer with CRISPR-based detection to improve the precision of lab-based, point-of-care, and field-deployable diagnostic and biosurveillance solutions.

We next evaluated the capabilities of LAMP-based OnePot DETECTR using MB-CasFire for analytical sensitivity, clinical sample detection, quantification, and multiplexing. We designed a OnePot DETECTR assay for SARS-CoV-2 detection based on our previously published RT-LAMP-assay that targets the N-gene (Broughton et al., 2020). A similar OnePot DETECTR assay was designed targeting RNase P POP7 that could serve as a sample control. The MB-CasFire gRNA binding site for both assays was placed between the F1c and B1c primer binding sites within the LAMP amplicon to minimize overlap with the six primers used in LAMP (Fig. 2a). In general, for LAMP-based OnePot DETECTR, we avoided designs in which the gRNA binding site overlapped a primer binding site or was located between the F3 and F2 or B3 and B2 primer binding regions.

**Figure 2.**
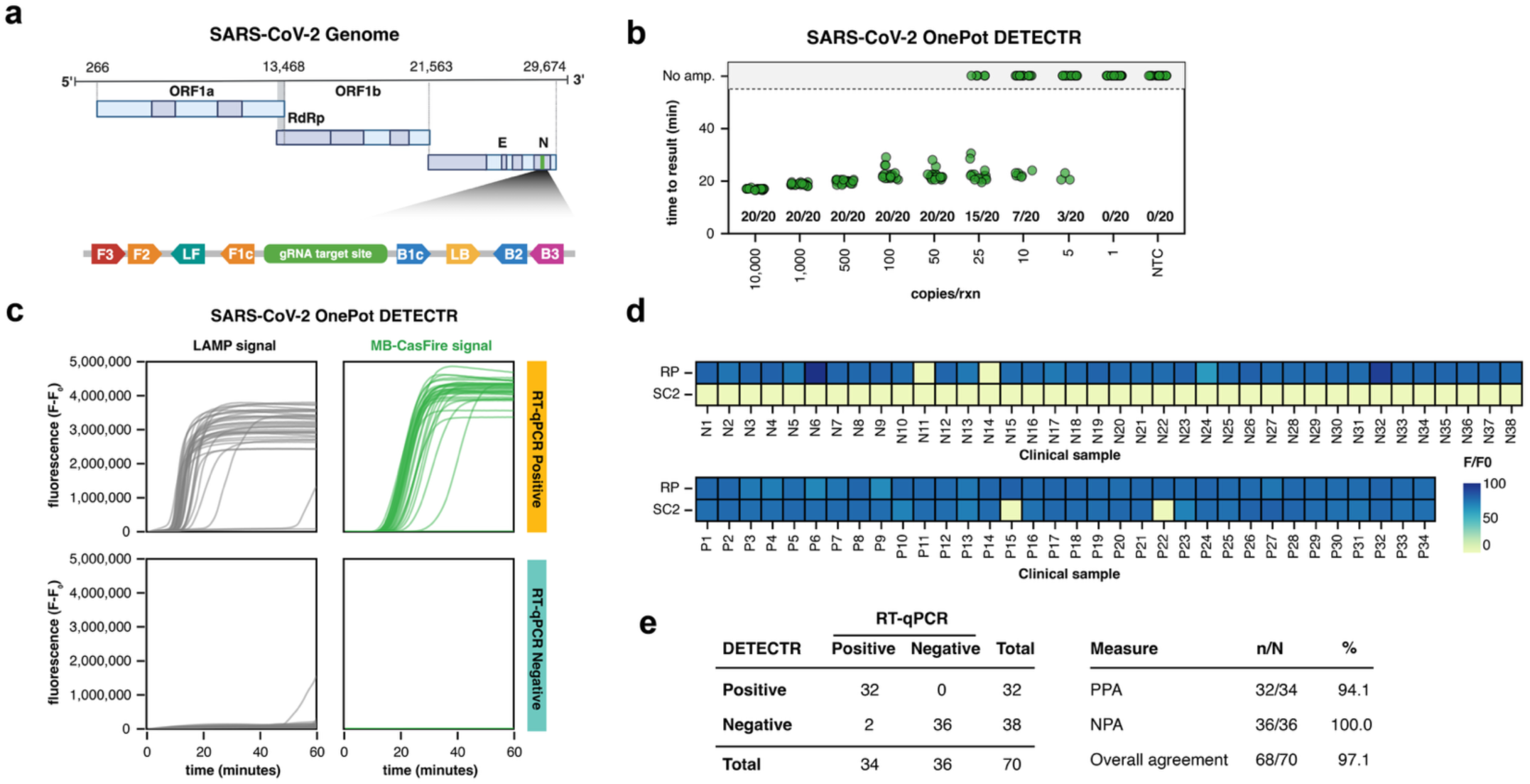
LAMP-based OnePot DETECTR using MB-CasFire for the detection of SARS-CoV-2. **(a)** Schematic of LAMP primer binding sites and position of gRNA target site on the N-gene of SARS-CoV-2. Created in BioRender. Broughton, J. (2026) https://BioRender.com/wp5o48q. **(b)** Analytical sensitivity of SARS-CoV-2 OnePot DETECTR. n=20 per input level. **(c)** Kinetic curves from SARS-CoV-2 OnePot DETECTR on clinical samples. Gray lines represent signal from DNA-binding dye monitoring LAMP amplification and green lines represent the signal from MB-CasFire. n=1 per clinical sample, 70 samples total. **(d)** Heatmap of end-point fluorescence (60 minutes) for MB-CasFire signal from OnePot DETECTR assays for SARS-CoV-2 (SC2) and human RNase P (RP), which served as a sample control. n=1 per clinical sample. **(e)** Table showing positive percent agreement (PPA) and negative percent agreement (NPA) of OnePot DETECTR compared to RT-qPCR assay for SARS-CoV-2 run on the same samples.

In the analytical sensitivity experiment, OnePot DETECTR detected all 20 replicates at 50 copies per reaction (cp/rxn), compared with 15 of 20 at 25 cp/rxn and 7 of 20 at 10 cp/rxn (Fig. 2b). We then tested RNA extracted from clinical nasopharyngeal (NP) swab samples with OnePot DETECTR and, in parallel, with an RT-qPCR comparator assay. Both LAMP and MB-CasFire produced strong fluorescent signal in SARS-CoV-2-positive samples. In one negative sample, non-specific amplification generated LAMP fluorescence but no MB-CasFire signal above background (Fig. 2c). The RNase P sample control was detected by MB-CasFire in all samples except two SARS-CoV-2-negative samples, both of which were also RNase P-negative by RT-qPCR (Fig. 2d). Excluding these two samples, OnePot DETECTR showed 94.1% positive percent agreement (PPA, 32 of 34) and 100% negative percent agreement (NPA, 36 of 36) with RT-qPCR (Fig. 2e).

To extend OnePot DETECTR beyond qualitative detection, we asked whether the assay could quantify RNA input similar to RT-qPCR. Previous CRISPR-based assays achieved absolute quantification by partitioning reactions into digital formats (Wu et al., 2021; Ding et al., 2021), whereas OnePot DETECTR reports amplification and detection in a single real-time reaction. We designed a OnePot DETECTR assay for KRAS mRNA and defined time to result as the time at which MB-CasFire fluorescence crossed a detection threshold. Time to result decreased linearly with log input across a seven-point 10-fold dilution series of in vitro-transcribed (IVT) KRAS mRNA (R² = 0.98), comparable to RT-qPCR Ct (R² = 0.97, Supplementary Fig. 3). We then used this standard curve to estimate KRAS mRNA copies in total RNA isolated from 15 human normal and tumor tissues. In general, OnePot DETECTR estimates were higher than RT-qPCR estimates, although both methods ranked kidney tumor and testis highest (Supplementary Fig. 3e). Together, these results show that OnePot DETECTR can achieve relative quantification of RNA over six orders of magnitude.

To detect a viral target and a human sample control in one reaction, we searched for a second Cas enzyme to pair with MB-CasFire. Cas12a and Cas13a are orthogonal CRISPR enzymes with distinct trans-nuclease activity preferences for ssDNA and RNA, respectively, and have been multiplexed in a single detection reaction following a separate preamplification step (Gootenberg et al., 2018). A previous one-pot LAMP multiplex paired SLK9, which cleaves both poly-C and poly-T reporters, with AacCas12b, which cleaves only poly-T (Pena et al., 2023). This multiplexing approach is limited because it cannot discriminate between a sample that is positive for the SLK9 target and negative for the AacCas12b target, as both the poly-T and poly-C reporters would be degraded by SLK9’s trans-nuclease activity. We reasoned that pairing MB-CasFire with a Cas13a that cleaves an RNA reporter would give proper discrimination between targets in a OnePot DETECTR assay, provided the Cas13a could recognize the DNA products of LAMP. We identified a naturally thermostable Cas13a, MB-CasDx10, that cleaved its RNA reporter in response to both RNA and ssDNA targets in Cas-only reactions (Fig. 3a). Because strand displacement during LAMP releases single-stranded DNA and yields stem-loop products with single-stranded loops (Notomi et al., 2000), we tested whether MB-CasDx10 could detect LAMP product directly. Both MB-CasDx10 and MB-CasFire were capable of detecting LAMP products generated in a separate preamplification reaction (Fig. 3b).

**Figure 3.**
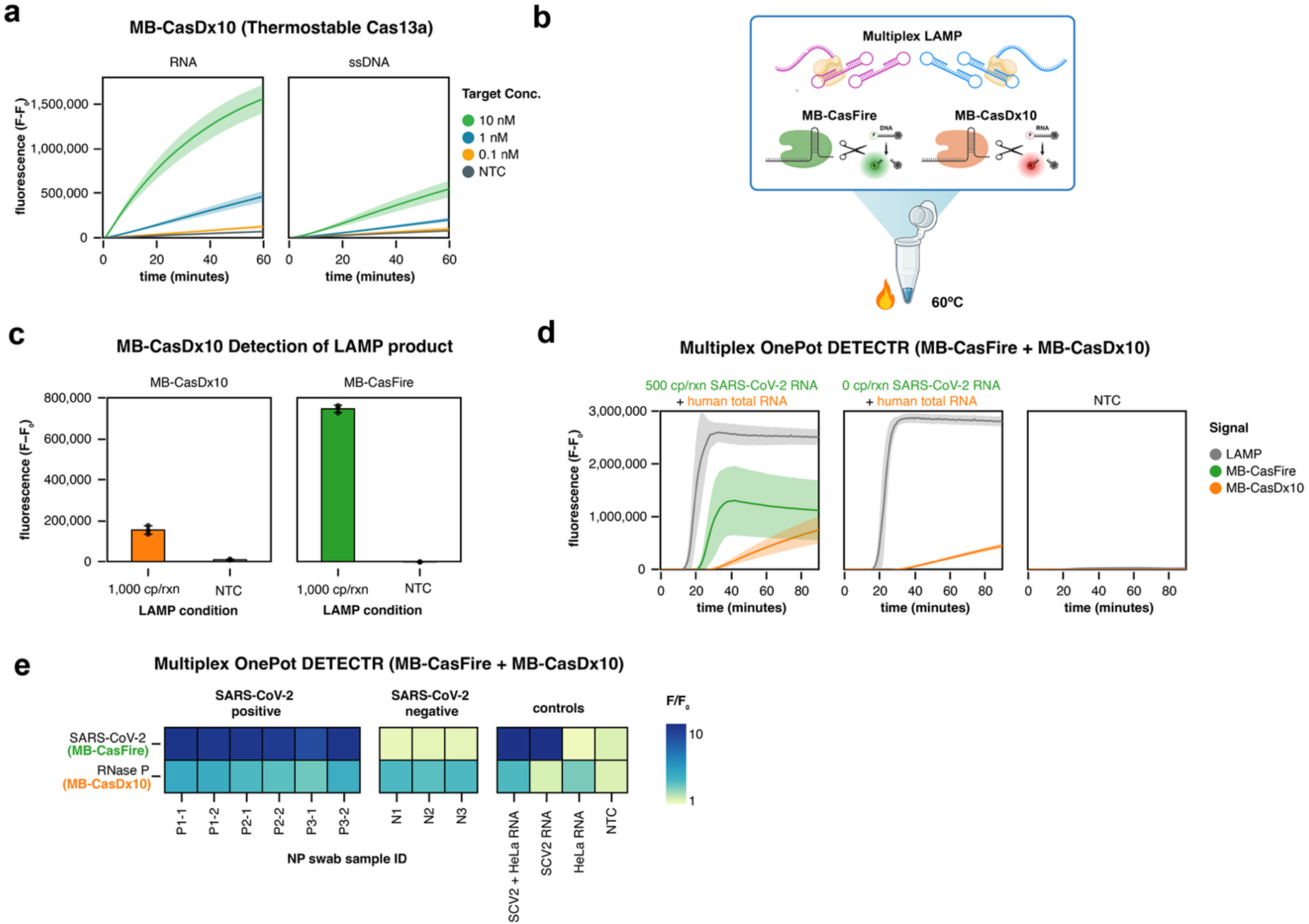
A thermostable Cas13a activated by ssDNA enables multiplexed OnePot DETECTR with MB-CasFire. **(a)** RNA reporter cleavage by MB-CasDx10 in Cas-only reactions with RNA or ssDNA target oligos at 10, 1, or 0.1 nM, or without target (NTC). Lines show the mean and shaded areas ± 1 SD (n = 3). **(b)** Schematic of multiplexed OnePot DETECTR. Created in BioRender. Broughton, J. (2026) https://BioRender.com/wp5o48q. **(c)** 30 minute endpoint fluorescence at 60°C from MB-CasDx10 (RNA reporter) and MB-CasFire (ssDNA reporter) detection of LAMP product amplified in a separate reaction from 1,000 cp/rxn input or from an NTC. Bars show the mean ± SD (n = 3).**(d)** LAMP (DNA-binding dye), MB-CasFire, and MB-CasDx10 signals in multiplexed OnePot DETECTR reactions containing 500 or 0 cp/rxn SARS-CoV-2 RNA with human total RNA, or in NTC reactions. Lines show the mean and shaded areas ± 1 SD (n =3). **(e)** Multiplexed OnePot DETECTR on RNA extracted from nasal swab specimens that were positive (P) or negative (N) for SARS-CoV-2 by RT-qPCR, and on control reactions containing synthetic SARS-CoV-2 RNA with HeLa total RNA, SARS-CoV-2 RNA alone, HeLa RNA alone, or no template (NTC). The heat map shows normalized fluorescence (F/F_0_) in the MB-CasFire (SARS-CoV-2) and MB-CasDx10 (RNase P) channels at 60 min.

We then combined both enzymes in a multiplexed OnePot DETECTR reaction (Fig. 3c). In contrast to other one-pot Cas13 assays, such as SHINE, that include T7 RNA polymerase and ribonucleotides to transcribe DNA amplicons into RNA (Arizti-Sanz et al., 2020), our multiplexed OnePot DETECTR assay leveraged MB-CasDx10’s activation by ssDNA to simplify the reaction components. The assay reported LAMP amplification with a DNA-binding dye, SARS-CoV-2 amplicon with MB-CasFire, and human RNase P amplicon with MB-CasDx10. In reactions containing human total RNA, the MB-CasFire channel became positive only when SARS-CoV-2 RNA was added, whereas the MB-CasDx10 channel was positive with or without viral RNA, and no channel produced signal in the NTC (Fig. 3d). The multiplexed assay also detected SARS-CoV-2 and human RNA in extracted clinical samples (Fig. 3e). In comparison to MB-CasFire, MB-CasDx10 had slower signal kinetics and a longer time to detection (Fig. 3d), suggesting that further engineering and optimization of MB-CasDx10 could improve its performance in multiplexed assays.

To extend OnePot DETECTR to another amplification chemistry, we coupled MB-CasFire with Nicking Enzyme Amplification Reaction (NEAR) for the detection of *Neisseria gonorrhoeae*. NEAR amplifies DNA at a constant temperature by combining a nicking endonuclease with a strand-displacing DNA polymerase (Van Ness et al., 2003). Because it generates amplification products within minutes, NEAR is well suited to rapid molecular diagnostics. Consistent with this speed, NEAR-based OnePot DETECTR had a time to detection of approximately 7 min, compared with approximately 31 min for LAMP-based OnePot DETECTR with the same gRNA, a roughly 4.4-fold reduction (Fig. 4a).

**Figure 4.**
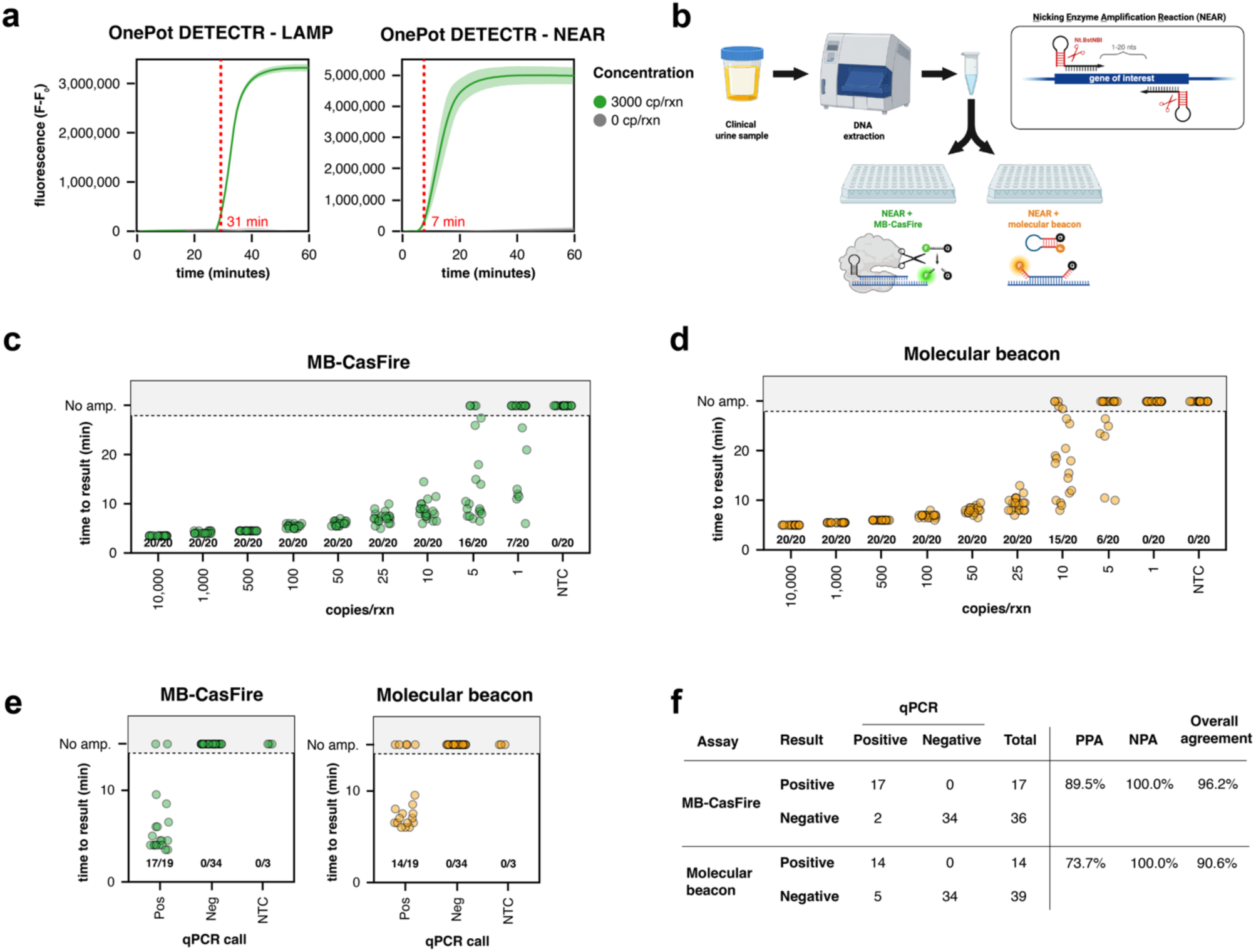
Comparison of a NEAR assay for N. gonorrhoeae using MB-CasFire and molecular beacons for readout. **(a)** Comparison of detection time of OnePot DETECTR assays using the same gRNA and either LAMP or NEAR as the isothermal amplification method. n=3, shaded area represents ± 1 SD. **(b)** Schematic of NEAR assays using either MB-CasFire or molecular beacon to detect N. gonorrhoeae from urine samples. Created in BioRender. Broughton, J. (2026) https://BioRender.com/wp5o48q. **(c)** Analytical sensitivity of NEAR-based OnePot DETECTR using MB-CasFire, n=20 per input level. **(d)** Analytical sensitivity of NEAR using a molecular beacon, n=20 per input level. **(e)** Time to result for the two assays across 19 reference-positive and 34 reference-negative clinical specimens, with three NTC reactions per assay. **(f)** Table summarizing the positive percent agreement (PPA) and negative percent agreement (NPA) of the two assays as compared to qPCR testing of the same extracted material.

NEAR assays are commonly read out with molecular beacons (Mozola et al., 2013), which are hairpin probes whose fluorophore and quencher separate when the probe hybridizes to its complementary amplicon (Tyagi and Kramer, 1996). We compared two NEAR assays using the same primers and buffer conditions, one was formulated as a OnePot DETECTR assay with MB-CasFire and the other used a molecular beacon (Fig. 4b). Both assays were run at 58°C. In titrations of genomic *N. gonorrhoeae* DNA spiked into pooled extract of normal urine, MB-CasFire detected inputs as low as 10 cp/rxn in 20 of 20 replicates, compared with 25 cp/rxn for the molecular beacon (Fig. 4c, d, Supplementary Fig. 4a, b). We then evaluated both NEAR assays on DNA extracted from 53 clinical urine specimens previously tested for *N. gonorrhoeae* by qPCR, of which 19 were positive and 34 negative. The MB-CasFire readout detected 17 of 19 reference-positive specimens (89.5% PPA), whereas the molecular beacon detected 14 of 19 (73.7% PPA). Both readouts correctly classified all 34 reference-negative specimens (100% NPA), giving overall agreement of 96.2% for MB-CasFire and 90.6% for the molecular beacon (Fig. 4f). With both readouts, positive calls came within 10 min, and MB-CasFire generally called positives earlier than the molecular beacon (Fig. 4e). Together, these results suggest that replacing a molecular beacon with MB-CasFire allows NEAR to detect lower target inputs and improves its performance on clinical samples.

We next assessed whether MB-CasFire could distinguish single-nucleotide variants, using a clinically relevant target in *N. gonorrhoeae*. The gyrA mutation encoding the S91F substitution is an important fluoroquinolone-resistance determinant associated with reduced ciprofloxacin susceptibility (Lee et al., 2021; Ison et al., 1998). To identify allele-specific gRNAs for MB-CasFire, we screened candidate gRNAs against synthetic DNA fragments carrying either the GyrA S91 or the S91F allele. We selected gRNA-S91 and gRNA-S91F, each of which preferentially detected its matched allele (Fig. 5a, b).

**Figure 5.**
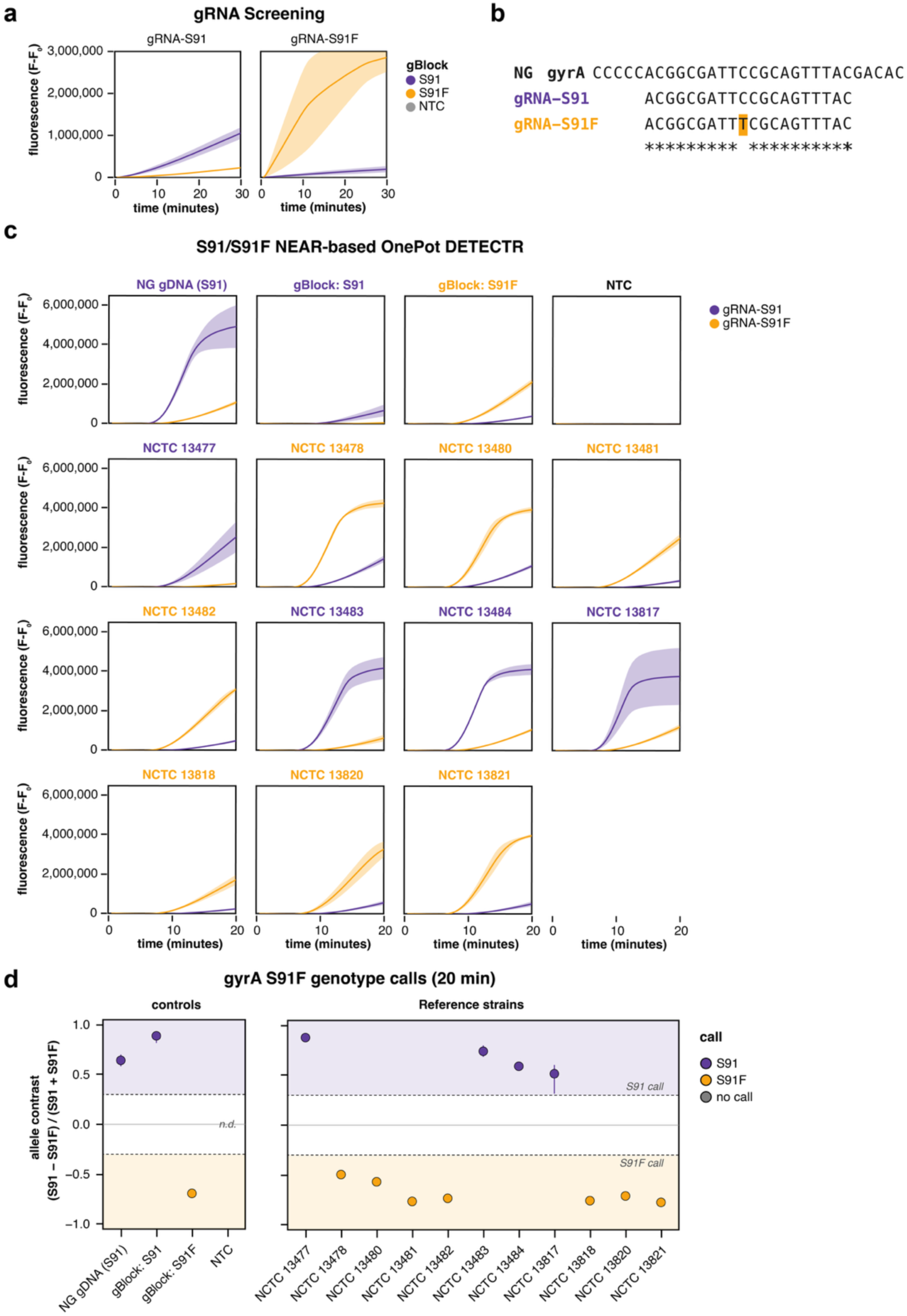
SNP-based detection using MB-CasFire for gyrA S91F in N. gonorrhoeae. **(a)** Kinetic curves from MB-CasFire detection assay on gBlocks containing either the sequence for S91 or S91F to identify SNP-sensitive gRNAs. n=3, shaded area represents ± 1 SD. **(b)** Alignment of SNP-sensitive guide spacer sequences against the wild-type gyrA sequence. **(c)** Evaluation of a NEAR-based OnePot DETECTR assay for the detection of S91/S91F. Each input is tested in separate reactions with gRNA-S91 or gRNA-S91F. DNA inputs were normalized to 3,000 cp/rxn, n=4, shaded area represents ± 1 SD. **(d)** gyrA S91F genotype calls at 20 min from the reactions in panel C. The allele contrast is calculated from background-subtracted fluorescence (F–F_0_) using the equation (S91 − S91F)/(S91 + S91F). Inputs with a contrast above 0.3 were called S91, those below −0.3 were called S91F, and those in between were not called (shaded regions). Points show the mean index, and error bars show the 95% confidence interval (n = 4). n.d., not detected.

We then evaluated allele discrimination by NEAR-based OnePot DETECTR using genomic DNA from 11 WHO *N. gonorrhoeae* reference strains with GyrA S91 or S91F genotypes (Unemo et al., 2016; Unemo et al., 2024; Fig. 5c). Each sample was tested with gRNA-S91 and gRNA-S91F, and genotypes were called by calculating the allele contrast from the MB-CasFire signal with each gRNA. Genotype calls at 20 min were 100% concordant (11 of 11) with the reference genotypes (Fig. 5d) and were confirmed by TaqMan genotyping. These results show that MB-CasFire can discriminate single-nucleotide variants within a OnePot DETECTR reaction, extending MB-CasFire from pathogen detection to genotyping of clinically relevant mutations.

## DISCUSSION

MB-CasFire was developed to enable sensitive and specific nucleic acid detection in a simplified workflow for point-of-care diagnostics and field-deployable biosurveillance solutions. These applications require an enzyme that is active at isothermal amplification temperatures, efficiently trans-cleaves the reporter within the amplification reaction, and selectively recognizes the target sequence. Starting from a naturally thermostable Cas12a, we improved performance under relevant molecular detection conditions and demonstrated applications in both LAMP- and NEAR-based OnePot DETECTR. The engineering of mesophilic MB-CasDx8 and MB-CasDx9 toward activity at elevated temperatures further expands the diagnostic enzyme portfolio. In a high-salt buffer, MB-CasFire exhibited the highest trans-nuclease activity rates of all the enzymes tested at each temperature from 50 to 75°C (Fig. 1d). Combining a Cas nuclease with isothermal amplification in a single reaction has been previously shown to reduce sensitivity and speed, because the nuclease cleaves the amplicon and thereby suppresses amplification (Tong et al. 2024). However, here we show that by engineering and optimizing CRISPR enzymes for OnePot DETECTR, we have developed MB-CasFire as a plug-and-play CRISPR detection nuclease that is compatible with multiple amplification chemistries, and unlocked new capabilities such as SNP discrimination in a rapid and simplified workflow.

MB-CasFire was compatible with both LAMP- and NEAR-based OnePot DETECTR, indicating flexibility in the choice of amplification chemistry during assay development (Fig. 2 and Fig. 4). Using the same gRNA, NEAR-based assays generally detected targets faster than LAMP-based assays (Fig. 4a), an advantage for applications that require rapid detection. MB-CasFire also improved detection and assay performance relative to a molecular beacon readout of a NEAR assay that used the same primers and buffer conditions (Fig. 4). We speculate that the improved performance of NEAR-based OnePot DETECTR reflects catalytic signal amplification by MB-CasFire. Once activated by a target, Cas nucleases such as Cas12a cleave many reporter molecules through multiple-turnover trans-nuclease activity (Chen et al., 2018), whereas a molecular beacon generates signal only while hybridized to an amplicon.

In addition to improving sensitivity, combining MB-CasFire with isothermal amplification in OnePot DETECTR improves assay specificity by adding a second recognition layer that eliminates detection of non-specific amplification (Fig. 2c). Isothermal amplification methods such as LAMP and RPA are prone to non-specific amplification, which can produce false-positive results (Li et al., 2019; Rolando et al., 2020). When amplification is read out with a DNA intercalating dye, which reports any double-stranded product, primers must be designed to avoid primer dimers, self-amplifying hairpins, and mispriming (Meagher et al., 2018; Hardinge and Murray, 2019). This requirement can leave few usable primer sets when sequence conservation restricts the target region, as in pan-influenza and pan-rhinovirus assays (Nakauchi et al., 2019). Because reporter cleavage by MB-CasFire requires gRNA recognition of the target sequence within the amplicon, MB-CasFire discriminates target-specific from non-specific amplification, which could enable primer designs that would otherwise be unusable. Beyond specificity, pairing MB-CasFire with MB-CasDx10 enabled multiplexed detection of SARS-CoV-2 and RNase P in a single RT-LAMP reaction (Fig. 3d). Cas13a-based detection of amplified DNA typically requires T7 transcription to convert the amplicon to RNA (Gootenberg et al., 2017). In contrast, MB-CasDx10 detected its target without T7 RNAP or other transcription components added to the reaction, consistent with direct activation of MB-CasDx10 by ssDNA generated during LAMP and with the DNA-activated collateral cleavage reported for LbuCas13a (Wu et al., 2025).

Single-nucleotide discrimination is typically not a capability of isothermal amplification methods because the assays can tolerate single-base mismatches between the primers and templates, as shown for RPA and LAMP (Daher et al., 2015; Varona and Anderson, 2021), and the reaction runs at a fixed temperature that cannot be raised to make primer or probe binding more stringent. Cas12a can also tolerate individual guide–target mismatches, and discrimination depends on mismatch position, nucleotide identity, and guide design (Kohabir et al., 2024), but appropriate design of gRNAs enables SNP detection by OnePot DETECTR. For example, previous NEAR-Cas12b and RPA-Cas12a platforms have addressed these constraints through guide engineering and control of amplification-detection kinetics (Bai et al., 2025; Wu et al., 2026). MB-CasFire demonstrated allele-specific detection of the *gyrA* S91F locus in both synthetic DNA templates and NEAR reactions with genomic reference material. Although the maximum signal varied between reference strains, the gRNA matching the expected genotype gave a concordant call in every strain tested (Fig. 5). Given the prospective clinical evidence supporting *gyrA* codon 91 genotyping to guide ciprofloxacin treatment (Klausner et al., 2021), this application is an example of how rapid detection of antibiotic resistance could enable actionable decision-making at the point-of-care.

Here we report the discovery of three novel Type V CRISPR enzymes with trans-nuclease activity from our metagenomic database, and subsequent protein engineering that led to improved thermostability and compatibility with several isothermal amplification chemistries. These results establish MB-CasFire as a robust, plug-and-play DETECTR enzyme that has the potential to combine the sensitivity and specificity with simplified workflows, and unlocks new capabilities such as improved speed and signal-to-noise for target detection in clinical samples, greater primer design flexibility, and SNP calling for genotyping applications. MB-CasFire is the latest tool in the CRISPR diagnostics portfolio, and its unique capabilities can enable inexpensive devices without compromising performance that could reshape the field of point-of-care molecular diagnostics and field-deployable biosurveillance applications.

## Data Availability

All data are available from the corresponding author upon reasonable request, subject to confidentiality obligations of Mammoth Biosciences, Inc.

## ACKNOWLEDGEMENTS

We thank New England Biolabs (NEB) for helpful discussions and critical review of the manuscript. Clinical urine samples that were used to evaluate the NEAR-based assays for *N. gonorrhoeae* porA were obtained from the laboratory of Dr. Benjamin Pinsky at Stanford University.

## SUPPLEMENTARY FIGURES

**Supplementary Figure 1.**
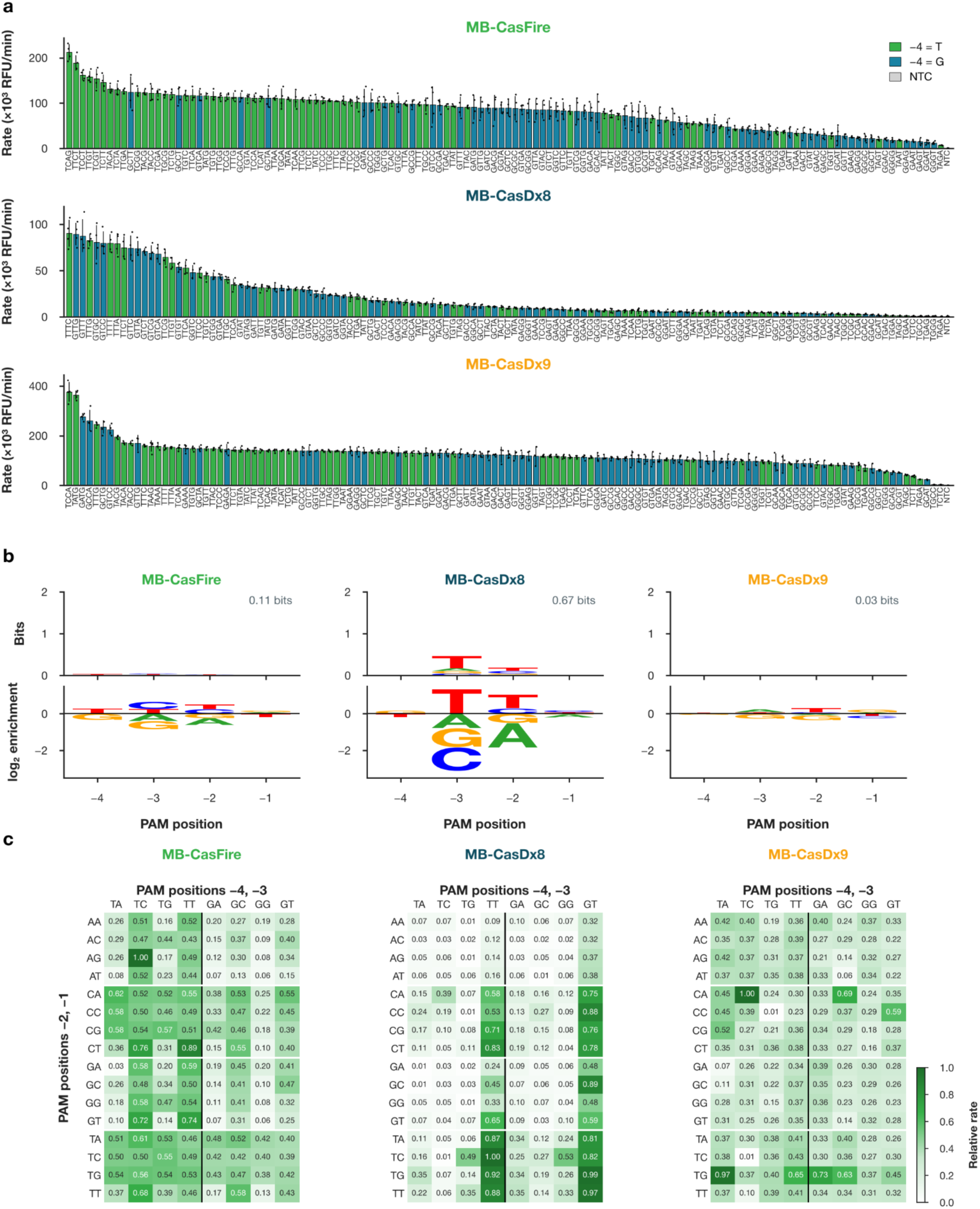
PAM preferences of MB-CasFire, MB-CasDx8, and MB-CasDx9. **(a)** Trans-nuclease activity rate of MB-CasFire, MB-CasDx8, and MB-CasDx9 against 128 double-stranded DNA targets carrying 5′-TNNN-3′ or 5′-GNNN-3′ protospacer adjacent motifs (PAMs). Bars are colored by the nucleotide at position -4 (T, green; G, teal). Error bars show the standard deviation. **(b)** Rate-weighted PAM logos for each enzyme. The top logo shows information content relative to the library composition, with total bits indicated. The bottom logo shows the log2 ratio of the mean NTC-subtracted rate of PAMs. Letters above zero mark nucleotides whose PAMs are faster than average, letters below zero mark slower PAMs, and a value of 1 corresponds to a twofold higher mean rate. Position -4 shows only T and G, the two nucleotides present in the library. **(c)** NTC-subtracted mean rate for each PAM normalized to the fastest PAM of the same enzyme, arranged by positions -4 and -3 (columns) and -2 and -1 (rows).

**Supplementary Figure 2.**
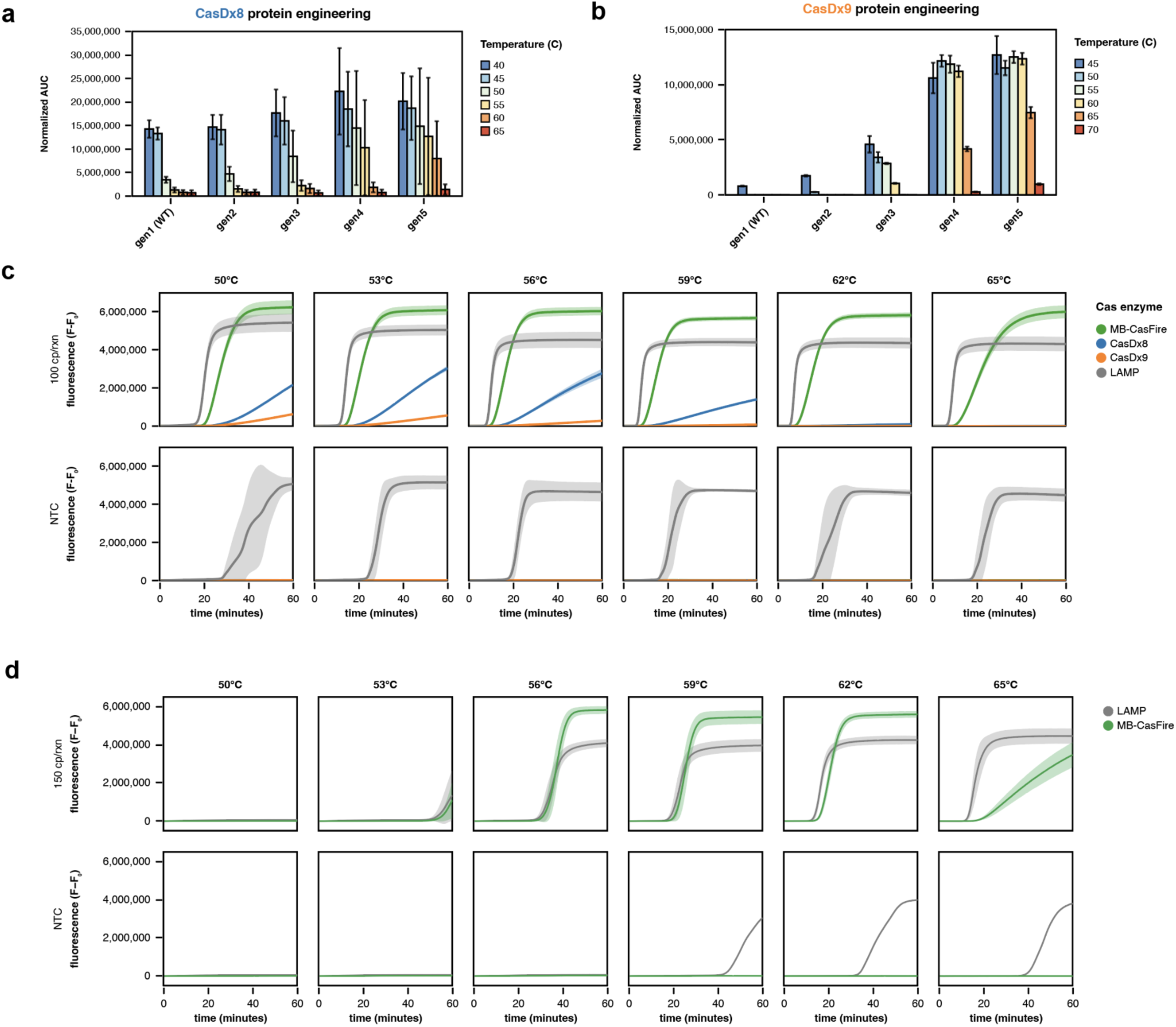
Protein engineering and temperature-dependent performance of diagnostic Cas enzymes. **(a, b)** Trans-nuclease activity across successive protein engineering generations of CasDx8 **(a)** and CasDx9 **(b)**, measured in an isothermal amplification buffer. Area under the fluorescence curve (AUC) is normalized to a non-target control. **(c)** Temperature-dependent performance of OnePot DETECTR reactions containing MB-CasFire (green), CasDx8 (blue), or CasDx9 (orange), with influenza B RNA at 100 copies per reaction (top) or no-template controls (NTCs; bottom). LAMP amplification is shown in gray. n=4. **(d)** Temperature-dependent amplification and detection kinetics for MB-CasFire-based OnePot DETECTR with SARS-CoV-2 at 150 copies per reaction (top) or NTCs (bottom). DNA amplification was monitored using a DNA-binding dye (gray), and MB-CasFire trans-nuclease activity was monitored using a fluorescent ssDNA reporter (green). Bars and curves represent mean, and error bars and shaded regions indicate +/- 1SD.

**Supplementary Figure 3.**
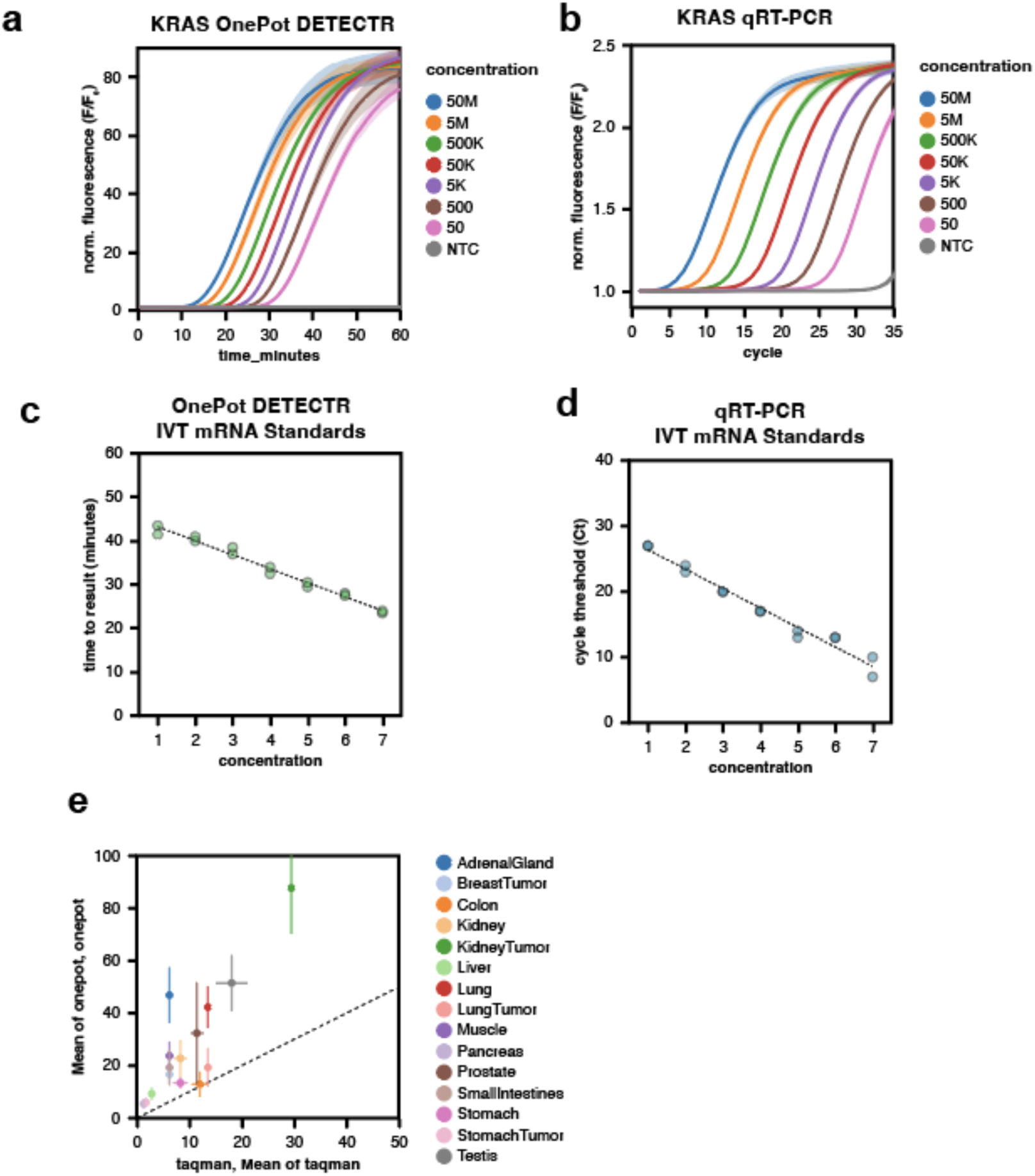
OnePot DETECTR quantifies KRAS mRNA across six orders of magnitude and returns higher estimates than qRT-PCR in human tissue RNA. **(a)** Normalized MB-CasFire reporter fluorescence (F/F_0_) during 60 min of OnePot DETECTR at 58°C with in vitro–transcribed (IVT) KRAS mRNA standards from 50 to 5 × 10⁷ copies per reaction and a no-template control (NTC). **(b)** Normalized TaqMan probe fluorescence (F/F_0_) over 35 cycles of qRT-PCR with the same standards. **(c)** OnePot DETECTR time to result for IVT standards at seven 10-fold dilution levels, numbered 1 (lowest) to 7 (highest), run on the same plate as the OnePot DETECTR tissue samples in e. **(d)** qRT-PCR cycle threshold (Ct) for the same dilution series, run on the same plate as the qRT-PCR tissue samples. **(e)** KRAS mRNA copies per reaction in total RNA from 15 human normal and tumor tissues, estimated by each method from the standard curves in c and d. F/F_0_ is fluorescence divided by the first reading of the same well. Lines and shaded bands in a and b show the mean ± SD of five replicate reactions from one operator. In c and d, points are individual reactions run in duplicate, and dashed lines are linear fits against dilution level (R² = 0.98 for OnePot DETECTR and 0.97 for qRT-PCR). In e, points show the mean and error bars the SEM of seven replicate reactions per tissue for each method, and the dashed line marks equal estimates (y = x). Time to result was when the reporter signal crossed the threshold, and Ct was the second-derivative maximum of ROX-normalized probe fluorescence.

**Supplementary Figure 4.**
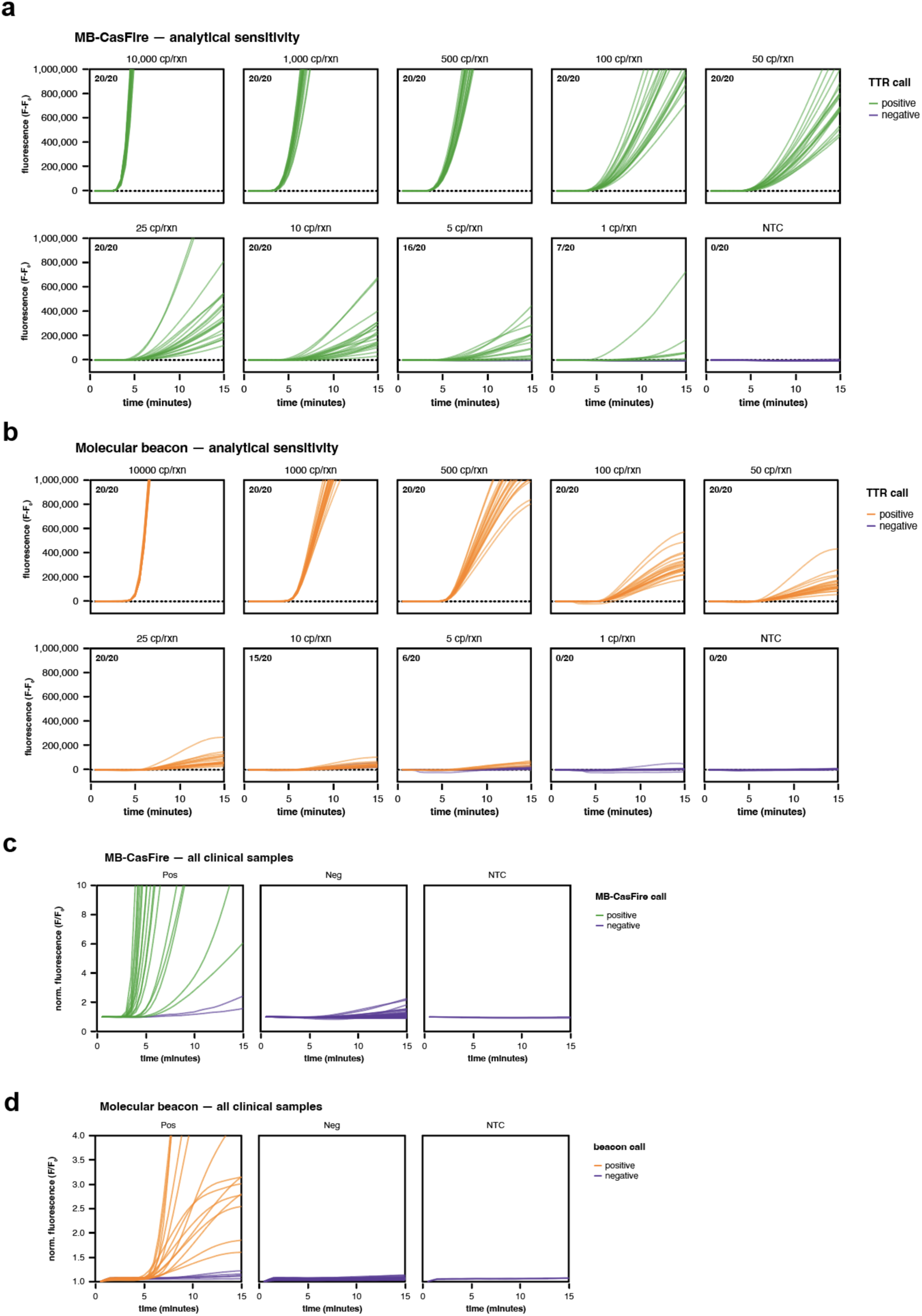
Individual kinetic curves for the evaluation of NEAR-based assays for N. gonorrhoeae using either MB-CasFire or a molecular beacon for a readout. **(a)** Analytical sensitivity of NEAR-based OnePot DETECTR. Replicates called positive are colored green, while replicates called negative are colored purple. **(b)** Analytical sensitivity of NEAR using a molecular beacon. Replicates called positive are colored orange, while replicates called negative are colored purple. **(c)** Clinical-sample fluorescence traces using NEAR-based OnePot DETECTR. Replicates called positive are colored green, while replicates called negative are colored purple. **(d)** Clinical-sample fluorescence traces using NEAR with a molecular beacon. Replicates called positive are colored orange, while replicates called negative are colored purple.

## MATERIALS & METHODS

### OnePot DETECTR for SARS-CoV-2 using LAMP

OnePot DETECTR reactions for SARS-CoV-2 combined RT-LAMP with MB-CasFire trans-nuclease activity in a single 10 µL reaction. MB-CasFire and gRNA were complexed at 400 nM each in 1X isothermal amplification buffer (Mammoth Biosciences) for 15 min at 37°C, and 1 µL of the ribonucleoprotein (RNP) complex was added per reaction to the RT-LAMP master mix for a final concentration of 40 nM. Each reaction contained 5 µL of WarmStart Multi-Purpose LAMP/RT-LAMP 2X Master Mix with UDG (NEB, M1708L), supplemented with 0.04 U/µL thermostable inorganic pyrophosphatase (NEB, M0296L). LAMP primers targeting the SARS-CoV-2 N gene (Broughton et al. 2020) were used at 0.2 µM F3 and B3, 1.6 µM FIP and BIP, and 0.8 µM LF and LB. The gRNA target site lies between the F1c and B1c primer binding sites of the amplicon (Fig. 2a). Amplification was monitored with 1 µM SYTO 9, and MB-CasFire trans-nuclease activity with 1 µM of CasDx ssDNA reporter labeled with Alexa Fluor 594 and Iowa Black RQ (IDT). Each reaction received 2 µL of quantified SARS-CoV-2 genomic RNA (ATCC VR-3276SD) diluted in nuclease-free water to 150 copies per reaction. Reactions were incubated for 60 min at 50, 53, 56, 59, 62, or 65°C on the gradient block of a QuantStudio 5 (Applied Biosystems). SYTO 9 fluorescence was read in the FAM channel and reporter fluorescence in the JUN channel.

### Multiplexed OnePot DETECTR for SARS-CoV-2 using LAMP

Multiplexed OnePot DETECTR combines MB-CasFire with MB-CasDx10 to recognize target ssDNA generated during a multiplexed LAMP assay. Each enzyme was complexed separately with its gRNA at a 1:1 molar ratio with a final concentration of 40 nM MB-CasFire RNP and 60 nM MB-CasDx10 RNP. Reactions (10 µL) contained isothermal reaction buffer (Mammoth Biosciences), 0.6 U/µL WarmStart Bst 2.0 DNA polymerase (NEB, M0538L), 0.375 U/µL WarmStart RTx reverse transcriptase (NEB, M0380L), 0.04 U/µL thermostable inorganic pyrophosphatase (NEB, M0296L), 0.2 U/µL murine RNase inhibitor (NEB, M0314L), 1 mM dNTPs (NEB, N0447L), and 500 nM SYTO 9. The SARS-CoV-2 N gene primer set was used at half the concentrations described above and was combined with a LAMP primer set targeting RNase P POP7. MB-CasFire cleavage was read with 750 nM of a Cy5-labeled poly(T) ssDNA reporter, and MB-CasDx10 cleavage with 1 µM of an Alexa Fluor 594-labeled poly(U) RNA reporter, both quenched with Iowa Black RQ (IDT). Reactions were incubated at 58°C for 60 or 90 min with fluorescence read in the FAM (SYTO 9), Cy5 (SARS-CoV-2), and JUN (RNase P) channels.

### OnePot DETECTR for *N. gonorrhoeae* porA using NEAR

NEAR-based OnePot DETECTR for *Neisseria gonorrhoeae* targeted the porA pseudogene with a single NEAR primer pair, each primer carrying a 5′ Nt.BstNBI recognition site and stabilizing region. MB-CasFire was complexed with gRNA at a 1:1 molar ratio for 20 min at 37°C, and the RNP was added to a final concentration of 40 nM. Reactions (10 µL) contained 0.75 U/µL WarmStart Nt.BstNBI (NEB, R0725S), 0.5 U/µL WarmStart Bst 2.0 DNA polymerase (NEB, M0538L), and 0.05 U/µL thermostable inorganic pyrophosphatase (NEB, M0296L) in a Tris-based NEAR buffer containing 7 mM MgSO4. Each reaction also contained 0.5 mM dNTPs (NEB, N0447L), 1 µM of each primer, and 1 µM of the CasDx reporter. Clinical urine (Stanford) or pooled normal human urine (Medix Biochemica) specimens were extracted on an EZ1 instrument with the EZ1 DSP Virus Kit (Qiagen), and eluted in 60 µL. Reactions were incubated at 58°C for 15 or 30 min on a QuantStudio 5 (Applied Biosystems), with reporter fluorescence read in the JUN channel at 30 sec intervals.

### Detection of *N. gonorrhoeae* porA using NEAR with a molecular beacon

For comparison with OnePot DETECTR, the same porA NEAR primers and urine extracts were tested with a molecular beacon readout. The beacon was a 22-nt probe complementary to the porA amplicon, with a 5-bp self-complementary stem, a 5′ 6-carboxyfluorescein (FAM) fluorophore, and a 3′ Iowa Black FQ quencher (IDT). Reactions (10 µL) contained 0.3 U/µL WarmStart Nt.BstNBI (NEB, R0725S) and 0.5 U/µL WarmStart Bst 2.0 DNA polymerase (NEB, M0538L) in the NEAR buffer described above. Each reaction also contained 0.5 mM dNTPs, 100 nM forward primer, 500 nM reverse primer, 200 nM beacon, and 2 µL of extract. Reactions were incubated at 58°C for 60 min on a QuantStudio 5 (Applied Biosystems), with FAM fluorescence read at 30 sec intervals.

### OnePot DETECTR for *N. gonorrhoeae* gyrA S91F using NEAR

To discriminate the gyrA S91F mutation, each sample was tested in parallel reactions containing one of the two gRNAs, with MB-CasFire RNP complexed at a 1:1 molar ratio for 15 min at 37°C and added to 40 nM. NEAR reactions (10 µL) contained 0.75 U/µL WarmStart Nt.BstNBI (NEB, R0725S), 0.5 U/µL WarmStart Bst 2.0 DNA polymerase (NEB, M0538L), and 0.05 U/µL thermostable inorganic pyrophosphatase (NEB, M0296L). Each reaction also contained 0.5 mM dNTPs (NEB, N0447L), 0.4 µM forward and 1.6 µM reverse primers, and 1 µM CasDx ssDNA reporter. Templates were genomic DNA from 11 N. gonorrhoeae NCTC reference strains, quantified genomic DNA (ATCC 700825DQ), and 700-bp synthetic gene fragments encoding S91 or F91. Genomic DNA copy number was calculated and added at a final concentration of 3,000 copies per reaction. Reactions were run at 58°C for 20 min on a QuantStudio 5 (Applied Biosystems) in 384-well plates, with reporter fluorescence read in the JUN channel. Genotyping calls were made based on the allele contrast, which was calculated from background-subtracted fluorescence (F–F_0_) using the equation (S91 − S91F)/(S91 + S91F).

### DETECTR-based PAM profiling

PAM preferences of MB-CasFire, MB-CasDx8, and MB-CasDx9 were profiled by trans-nuclease activity against a clonal library of 128 double-stranded DNA (dsDNA) targets. Each target carried a 4-nt PAM immediately 5′ of a shared 20-nt protospacer, with position −4 fixed as T or G and positions −3 to −1 covering all 64 combinations. Targets were PCR-amplified from clonal plasmids with Q5 High-Fidelity 2X Master Mix (NEB, M0492L), purified with SPRI beads, and normalized to 100 nM. Each enzyme was complexed with its cognate guide RNA (20 nM each) in 1X IB1 buffer (Mammoth Biosciences) for 15 min at 37°C. A 6-FAM-labeled poly(T)12 single-stranded DNA (ssDNA) reporter quenched with Iowa Black FQ (IDT) was then added to 200 nM. Reactions (10 µL) containing 100 pM target were run in 384-well plates with four replicate wells per PAM, and each plate included a randomized-PAM target (NNNN) and a no-target control (NTC). Reactions were incubated at 55°C (MB-CasFire, MB-CasDx9) or 50°C (MB-CasDx8) on a QuantStudio 5 (Applied Biosystems), with FAM fluorescence read every 30 sec for 30 min. The trans-nuclease activity rate of each well was the maximum slope of a 2.5-min sliding linear fit, and wells whose first read indicated a dispensing failure were excluded. Rates were averaged across wells, corrected by subtracting the plate NTC rate, and displayed as rate-weighted sequence logos generated with Logomaker (Tareen and Kinney, 2020).

